# Generation of realistic synthetic data using multimodal neural ordinary differential equations

**DOI:** 10.1101/2021.09.26.21263968

**Authors:** Philipp Wendland, Colin Birkenbihl, Marc Gomez-Freixa, Meemansa Sood, Maik Kschischo, Holger Fröhlich

## Abstract

Individual organizations, such as hospitals, pharmaceutical companies and health insurance providers are currently limited in their ability to collect data that is fully representative of a disease population. This can in turn negatively impact the generalization ability of statistical models and scientific insights. However, sharing data across different organizations is highly restricted by legal regulations. While federated data access concepts exist, they are technically and organizationally difficult to realize. An alternative approach would be to exchange synthetic patient data instead. In this work, we introduce the Multimodal Neural Ordinary Differential Equations (MultiNODEs), a hybrid, multimodal AI approach, which allows for generating highly realistic synthetic patient trajectories on a continuous time scale, hence enabling smooth interpolation and extrapolation of clinical studies. Our proposed method can integrate both static and longitudinal data, and implicitly handles missing values. We demonstrate the capabilities of MultiNODEs by applying them to real patient-level data from two independent clinical studies and simulated epidemiological data of an infectious disease.

## Introduction

Patient-level data build the foundation for a plethora of healthcare research endeavors such as drug discovery, clinical trials, biomarker discovery, and precision medicine [1]. Collecting such data is extremely time-consuming and cost-intensive, and additionally access-restricted by ethical and legal regulations in most countries. Individual organizations, such as hospitals, pharmaceutical companies and health insurance providers are currently limited in their ability to collect data that is fully representative of a disease population. This issue is especially pronounced in clinical studies, where patients are usually recruited based on predefined inclusion and exclusion criteria which introduce cohort-specific statistical biases [2]. These biases, in turn, can negatively impact the generalization ability of machine learning models, since the usual i.i.d. assumption is violated [3]. A naïve idea to counteract this issue might be to build up large data repositories pooling diverse clinical studies from several organizations. However, here, a major obstacle is that sharing patient-level data across different organizations is exceedingly difficult due to legal restrictions, as formulated, for example, in the General Data Protection Rule (GDPR) of the European Union.

The idea we propagate in this paper is to learn a continuous-time generative machine learning model from clinical study data. Given the distribution of the real training data was appropriately learned by such a model, the generated synthetic datasets maintain the real data signals, such as variable interdependencies and time-dependent trajectories. Furthermore, these synthetic datasets can overcome crucial limitations of their real counterparts like missing values or irregular assessment intervals, hence opening the opportunity to make at least subsets of variables from different studies statistically comparable. A further strong motivation for generating synthetic datasets is the aim to use the generated data as an anonymized version of its real-world counterpart and thereby mitigate the increased restrictions for sharing human data [4, 5, 6]. However, synthetic patient-level datasets open opportunities that reach far beyond data sharing. For example, trained generative models could be used for synthesizing control arms for clinical trials based on data from previously conducted trials, or from real-world clinical routine data [7]. This helps addressing major ethical concerns in disease areas, such as cancer, where it is impossible to leave patients untreated. Both, the American Food and Drug Administration (FDA) and the European Medicines Agency (EMA) have recognized this issue and taken initiatives to allow for synthetic control arms [7].

Over the last years, generative models (mostly Generative Adversarial Networks - GANs) have found notable success, mostly in the medical imaging domain [8,9,19-22]. However, GANs are often found to show a collapse to the statistical mode of a distribution, which raises concerns regarding coverage of the real patient distribution by synthetic data. Moreover, these methods are not necessarily suited to cope with the complex nature of clinical data collected in observational, longitudinal cohort studies, which is the main focus of our work: In addition to the previously mentioned issue of irregular measurement frequencies and missing values not at random (for example due to participant drop-out), clinical studies often comprise several modalities combining time-dependent variables (e.g., measures of disease severity) and static information (e.g., biological sex). One approach specifically designed for the joint modeling and generation of multimodal, time-dependent and static patient-level data containing missing values are the recently introduced Variational Autoencoder Modular Bayesian networks (VAMBN) [4]. However, VAMBN only operates on a discrete time scale while relevant clinical indicators such as, for example, disease progression expressed through cognitive decline or rising inflammatory markers, are intrinsically time continuous. Recently, Neural Ordinary Differential Equations (NODEs) have been introduced as a hybrid approach fusing neural networks and ordinary differential equations (ODE) [10]. While NODEs are time continuous and thus enable smooth interpolation between observed data points and extrapolation beyond the observations in the data, they are not able to integrate static variables.

In this work, we present the Multimodal Neural Ordinary Differential Equations (MultiNODEs) as an extension of the NODEs. MultiNODEs allow learning a generative model from multimodal longitudinal and static data that may contain missing values not at random. To demonstrate MultiNODEs’ generative capabilities, we applied the model to clinical, patient-level data from an observational Parkinson’s disease (PD) cohort study (the Parkinson’s Progression Markers Initiative, PPMI [11]) and, additionally, a longitudinal Alzheimer’s disease (AD) data collection (National Alzheimer’s Coordination Center, NACC [12]). We compared the generated trajectories and correlation structure with the real counterpart. In this context, we additionally evaluated MultiNODEs’ performance against the previously published VAMBN approach. Furthermore, we assessed MultiNODEs’ interpolation and extrapolation performance. Finally, we investigated the influence of sample size, noisiness of the data, and longitudinal assessment density on the training of MultiNODEs in a systematic benchmark on data simulated from a mathematical model well-known in the epidemiology field.

## Results

### Conceptual Introduction of the MultiNODEs

MultiNODEs represent an extension of the original NODEs framework [10] that overcomes the limitations of its predecessor such that an application to incomplete datasets consisting of both static and time-dependent variables becomes feasible. Conceptually, MultiNODEs build on three key components **(Figure 1)**: 1) latent NODEs, 2) a variational autoencoder (more specifically a Heterogenous Incomplete Variational Autoencoder - HI-VAE - designed to handle multimodal data with missing values [13]), and 3) an implicit imputation layer [14]. The latent NODEs enable the learning and subsequent generation of continuous longitudinal variable trajectories. The longitudinal properties of the initial condition (i.e., the starting point for the ODE system solver of the latent NODEs) are defined by the output of a recurrent variational encoder which embeds the longitudinal input data into a latent space **(Figure 1 orange box)**. To allow for an additional influence of static variables on the estimation of the longitudinal variable trajectories, the second component, a HI-VAE, is introduced **(Figure 1 blue box)**. This component transforms the static information into a distinct latent space and the resulting embedding is used to augment the latent starting condition of the NODEs by concatenating the static variable embedding and the latent representation of the longitudinal variables **(Figure 1 ‘augmentation’)**. The HI-VAE component itself holds generative properties and conducts the synthesis of the static variables when MultiNODEs are applied in a generative setting. Conclusively, MultiNODEs integrate static variables (e.g., biological sex or genotype information) both to inform the learning of longitudinal trajectories, and in the generative process. Finally, to mitigate the original NODEs’ incapability of dealing with missing values, we introduced the imputation layer which implicitly replaces missing values during model training with learned estimates **(Figure 1 green box)**. For further details on the model architecture, training, and hyperparameter optimization, we refer to the Method section and Supplements, respectively.

**Figure 1:** Conceptual framework of MultiNODEs. Blue box: HI-VAE for the encoding and generation of static variables. Orange box: NODEs that learn and generate longitudinal trajectories. Green box: The imputation layer which can handle missing data implicitly during model training.

### Synthetic Data Generation using MultiNODEs

Generating synthetic data using MultiNODEs starts by randomly sampling a latent representation for both the static and longitudinal variables, respectively. The longitudinal variables in data space are then generated by first constructing the initial conditions of the latent ODE system (i.e., concatenating the static latent representation to the longitudinal one), followed by solving the ODE system given these initial conditions, and finally by decoding the result into data space. The static variables are generated by directly transforming their sampled latent representation into data space using the HI-VAE decoder.

MultiNODEs support two different approaches for the initial sampling of the latent representations, namely sampling from the prior distribution employed during model training and sampling from the learned posterior distribution of the input data.

During the posterior sampling procedure, the reparameterization trick [15] is applied to draw a latent representation from the posterior distribution learned from the training data. The amount of noise added in this process can be tuned, whereas greater noise will lead to a wider spread of the generated marginal distributions of the synthetic data. Alternatively, the latent representations can be sampled from the prior distributions imposed on the latent space during variational model training. We ensure statistical dependence between static and longitudinal variables by drawing their values from a Bayesian network that connects both latent representations such that the longitudinal variables are conditionally dependent on the static variables. More detailed descriptions of both generation procedures are provided in the Method section.

### Application cases: Parkinson’s disease and Alzheimer’s disease

We applied MultiNODEs to longitudinal, multimodal data from two independent clinical datasets with the goal of generating realistic synthetic datasets that maintain the real data properties. Details about the data pre-processing steps are described in the Supplementary Material.

The first dataset was the Parkinson’s Progression Markers Initiative (PPMI), an observational clinical study containing 354 de-novo PD patients who participated in a range of clinical, neurological, and demographic assessments which form the variables of the dataset. In total, a set of 25 longitudinal and 43 static variables was investigated.

Furthermore, as a second example, we applied MultiNODEs to longitudinal, multimodal data from the National Alzheimer’s Coordinating Center (NACC). NACC is a database storing patient-level AD data collected across multiple memory clinics. After preprocessing, the dataset used in this study contained 2284 patients and a set of 3 longitudinal and 4 static variables was investigated.

In the following sections, we will focus on the results achieved on the PPMI data and refer to the equivalent experiments based on the NACC data which are presented in the Supplementary Material.

### MultiNODEs generate realistic synthetic patient-level datasets

We applied prior as well as posterior sampling for comparison purposes. With each method, we generated the same number of synthetic patients as encountered in the real dataset to allow for a fair comparison. To assess whether the generated data followed the real data characteristics, we conducted thorough comparisons of the marginal distributions using qualitative, visual assessments and further, quantitatively compared the Jensen-Shannon divergence (JS-divergence) between the generated data and real distributions. The JS-divergence is bound between 0 and 1 with 0 indicating equal distributions. Additionally, we investigated the underlying correlation structure of the measured variables. Finally, we trained a machine learning classifier (Random Forest) that evaluated whether real and synthetic patients showed similar clinical characteristics when compared to real healthy control individuals from their respective studies. Across all these aspects, we evaluated MultiNODEs’ performance in comparison to the previously published VAMBN approach [4].

The synthetic data generated using MultiNODE generally exhibited marginal distributions which bore high similarity to their corresponding real counterparts **(Figure 2, Supplementary Table 1, Supplementary Figure 1**; equivalent figures for the NACC data are presented in **Supplementary Figure 4.)**. The average JS-divergences between the real and synthetic distributions calculated across all variables and timepoints amounted to 0.018 ± 0.015 and 0.011 ± 0.009 for the PPMI data generated from the prior and posterior, respectively. For NACC the average JS-divergence was 0.071 ± 0.055 and 0.029 ± 0.031 for prior and posterior sampling, respectively. With respect to PPMI, data generation from the posterior distribution resulted in synthetic data that resembled the real data significantly closer than those generated from the prior distribution (Mann-Whitney-U-test, p < 0.02).

**Figure 2:** Marginal distributions of real and synthesized data for multiple variables. Mean, standard deviation and KL-Divergence for the displayed variables can be found in **Supplementary Table 1**. Equivalent results for the NACC data are presented in **Supplementary Figure 5. A**, time dependent variable ‘SCOPA’ at month 12. **B**, time dependent variable ‘UPDRS2’ at month 24. **C**, static variable ‘Aβ.42’. **D**, categorical static variable ‘Handedness’.

Compared to VAMBN, the prior sampling method seemed to be inferior with respect to the average JS-divergence when using NACC (U-test, p = 0.038). However, no statistically significant difference in the performance of VAMBN compared to MultiNODE’s posterior sampling could be observed (U-test, p = 0.80). For PPMI, no significant differences were found between VAMBN and any of MultiNODEs’ generation approaches (U-test, p = 0.31 for the prior approach; U-test, p = 0.24 for the posterior).

In order to evaluate whether MultiNODEs learned not only to reproduce univariate distributions but actually captured their interdependencies accurately, we compared the correlation structure of the generated data to that of the real variables. Visualizations of the Spearman rank correlation coefficients showed that both the prior and posterior sampling generated synthetic data which successfully reproduced the real variables’ interdependencies **(Figure 3)**. Comparing the results against VAMBN generated data revealed that both generation procedures of MultiNODEs were significantly better at reproducing the real data characteristics: the Frobenius norm of real data correlation matrix resulted in 45.3, and with a Frobenius norm of 25.66 the VAMBN generated data placed substantially further from the real data than the MultiNODEs approaches with 62.63 and 56.47 for the prior and posterior sampling, respectively. This shows that MultiNODEs slightly overestimated the present correlations, while VAMBN underestimated them. Concordantly, the relative error (i.e., the deviation of the respective synthetic dataset’s correlation matrix from the real one normalized by the norm of the real correlation matrix), was 0.81, 0.62, and 0.46 respectively for VAMBN and MultiNODEs’ prior and posterior sampling, leaving MultiNODEs with a substantially lower error than the VAMBN approach.

**Figure 3:** Correlation structure of real and synthetic data expressed as spearman rank correlation coefficients. Equivalent results for the NACC data are shown in **Supplementary Figure 6. A**, real data. **B**, posterior sampling from MultiNODEs. **C**, prior sampling from MultiNODEs. **D**, VAMBN generated data.

### Assessment of the utility of generated synthetic patients for machine learning

To evaluate whether the generated synthetic patients could be reliably used in a machine learning context, we built a Random Forest classifier that aimed to distinguish between healthy individuals and diseased patients. The classifier was trained within a five-fold cross-validation scheme once using real and once using synthetic diseased patients. Additionally, we trained a classifier on each respective synthetic dataset (comprising synthetically generated diseased and healthy subjects) and evaluated their performance on the real data **(Table 1)**. As predictors, we used clinical symptoms and genetic markers that are characteristic for the disease in question. For PD (PPMI), these were the UPDRS scores which describe a series of motor and non-motor symptoms commonly encountered in PD patients, for AD (NACC), we predominantly used cognitive assessments and a genetic risk factor. Technical details about the classifiers can be found in the Supplementary Material. Distinguishing real patients from healthy control subjects was possible with a 10 times repeated five-fold cross validated performance of 0.97 ± 0.02 area under the receiver operator curve (AUC) and 0.90 ± 0.01 AUC for PPMI and NACC, respectively. On PPMI, all evaluated generative methods achieved almost equal performance, indicating that clinical characteristics of synthetic patients followed the same patterns as in real patients. In addition, the most relevant features were the same across the real and all synthetic data-trained classifiers **(Supplementary Figure 13)**.

**Table 1:** Performance (AUC) of machine learning classifiers differentiating between real healthy control subject and real as well as synthetic patients, respectively. Values represent the average and standard deviation across a 10-times repeated 5-fold cross-validation.

For NACC, some deviations were found between a classifier’s cross-validated performance on real data and the synthetic-data-based performances. Here, MultiNODEs’ posterior and VAMBN showed similar deviations in opposite directions, with the posterior slightly overperforming and VAMBN slightly underperforming. The performance on the data generated via MultiNODE’s prior sampling method deviated the most **(Table 1)**. When trained on synthetic data and evaluated on real data, all trained classifiers underperformed compared to classifiers trained on real data. The feature importances of predictors were highly similar between the real data-trained and the respective synthetic data-trained classifiers.

### Generating data in continuous time through smooth interpolation and extrapolation

One particular strength of MultiNODEs, that sets it apart from alternative approaches such as VAMBN, is its ability to model variable trajectories in continuous time. The latent ODE system allows for estimation of variable trajectories at any arbitrary time point and thereby opens possibilities for 1) the generation of smooth trajectories, 2) overcoming panel-data limitations through interpolation, and finally 3) extrapolation beyond the time span covered in training data themselves. Again, we evaluated these capabilities based on the PPMI and NACC datasets (for brevity, NACC results are presented in the Supplementary Material). For the following we only focused on the MultiNODE posterior sampling approach to generate synthetic subjects.

Comparing the median trajectories of variables from the real data to those generated using MultiNODEs revealed that MultiNODEs accurately learned and reproduced the longitudinal dynamics exhibited in the real data **(Figure 4)**. Generation from both the prior and posterior distribution led to synthesized median trajectories that closely resembled the real median trajectories. Equivalently, also the 97.5% and 2.5% quantiles of the synthetic data approximated the corresponding real quantiles closely, indicating a realistic distribution of the synthetic data across the observed time points. This observation held true for most of the time-dependent variables (plots for all variables are linked in the Supplementary Material).

**Figure 4:** Comparison of median trajectories including the 2.5% / 97.5% quantiles of longitudinal variables from synthetic and real PPMI data. Additional examples are provided in **Supplementary Figure 2**. A corresponding example for the NACC dataset is shown in **Supplementary Figure 7. A, B, C, D**, depict different longitudinal variables from the PPMI dataset.

We further assessed the interpolation and extrapolation capabilities of MultiNODEs. For interpolation, one time point was excluded from model training and subsequently data was generated for all time points including the one left out. Contrasting the interpolated / imputed values against the corresponding real values showed that MultiNODEs accurately reproduced the longitudinal dynamics of a variable, even for unobserved time points **(Figure 5 A, C)**. In this context, we further compared the interpolated values against synthetic data that was generated based on the complete real data trajectory. We observed that the mean JS-divergence calculated across all variables between the interpolated data and the real data was slightly higher (0.025 ± 0.011) than that of the real data and the synthetic data generated after training MultiNODEs on the complete trajectory (0.016 ± 0.011). Similarly, the relative error between the interpolated correlation matrix and the real data was again only marginally higher than between the complete data and the real data (0.48 and 0.46, respectively; **Supplementary Figure 4**).

**Figure 5:** Time-continuous interpolation and extrapolation of exemplary PPMI variables. The black box indicates the interpolated and extrapolated sections. Plots for additional variables are presented in **Supplementary Figure 3**. A corresponding example for the NACC dataset is shown in **Supplementary Figure 8. A**, interpolation of the UPDRS1 variable at month 24. **B**, extrapolation of the last five assessments of the UPDRS1 variable. **C**, distribution of the interpolated values for UPDRS1 at visit 24. **D**, distribution of the extrapolated values for UPDRS1 at month 42.

In order to test MultiNODEs’ extrapolation capabilities, only the first 24 months of assessment follow-up and the static variables were used during model training. The trained model was then applied to generate data for the remaining, left out time points of the longitudinal variables. In this course, 77 values were extrapolated while not every variable had the same number of follow-up assessments after month 24. Comparing the extrapolated synthetic data to the left out real data demonstrated reliable extrapolation beyond the training data **(Figure 5 B, D)**. As in the interpolation setting, we also compared the average JS-divergence between the extrapolated data and the real data with that between the real data and synthetic data that were generated after training MultiNODEs on the complete trajectory. As expected, we could see a larger difference between the JS-divergences compared to the interpolation setting with 0.037 ± 0.024 for the extrapolated data and 0.016 ± 0.009 for the synthetic data based on the complete trajectory. The correlation structure in the extrapolation culminated in a relative error of 0.64 compared to 0.46 when using the complete trajectory for training MultiNODEs (**Supplementary Figure 4**).

In addition, the marginal distributions at both the interpolated and extrapolated time points also followed those of the real data **(Figure 5 C, D)**.

### Systematic model benchmarking on simulated data

To explore the learning properties of MultiNODEs more systematically, we investigated how alternating training conditions with respect to measurement frequency, sample size, and noisiness of the data influence MultiNODEs’ generative performance.

The benchmarking data was simulated via the well-established Susceptible-Infected-Removed (SIR) model which is often used to describe the spread of infectious diseases and follows a highly non-linear structure: Let *S* (*t*) be the number of susceptible individuals at a timepoint *t, I* (*t*) be the number of infectious individuals at a timepoint *t* and *R* (*t*) be the number of removed or recovered individuals at a timepoint *t*. With *β* as transmission rate, *γ* as mean recovery / death rate and *N* =*S* (*t*)+ *I* (*t*)+ *R* (*t*) as fixed population size the SIR model can be defined by the ODE system presented in Equation (1):

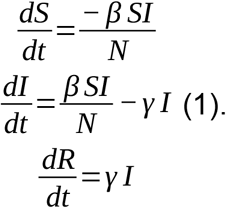

Details about the SIR parameter settings are described in the supplementary material.

As baseline settings for each investigation, we simulated 1000 data points with 10 equidistant assessment time points each, distributed over a span of 40 time intervals and added 5% Gaussian noise to each measurement. That means we added a normally distributed variable with the standard deviation set to 5% of the theoretical range of each of the variables S(t), I(t) and R(t). During the benchmarking, we individually alternated the sample size, time points and the noise level. For the time point investigation, we compared MultiNODEs’ trained on 5, 10 and 100 equidistant assessments; for the sample size we considered 100, 1000, and 5000 samples; and for the noise level we tested 50%, 75% and 100% of the maximum encountered value added as noise.

Alternating the amount of equidistant, longitudinal time points exposed a strong dependency of MultiNODEs on the longitudinal coverage of the time dependent process **(Figure 6 A)**. While the general trends in the data were appropriately learned for all explored assessment frequencies, the position of the observations in time influenced how close the learned function approximated the true data underlying process. Especially the peak of the ‘Infected’-function represented a challenge for MultiNODEs if no data point was located close to it **(Figure 6 A ‘Infected’)**. Similarly, the start of the decline in the ‘Susceptible’-function and the incline in the ‘Removed’-function were shifted, depending on the positioning of measurements. In conclusion, and as expected, a higher observation frequency of the data underlying time-dependent process significantly increased the fit of MultiNODEs to the process, although, general trends could already be approximated for lower assessment frequencies.

**Figure 6:** Model benchmarking on simulated data from the SIR-model. Each panel A, B, C represents the evaluation of another parameter (assessment frequency, sample size, noise level).

Investigating the effect of the sample size on training MultiNODEs, we observed that an increase of the sample size led to an expected improvement of the model fit to the SIR dynamics **(Figure 6 B)**. While the general trends could again be learned from limited data (n = 100), sample sizes of 1000 or 5000 substantially reduced the model’s deviation from the true SIR model. With 1000 samples, the learned dynamic is less stable than when trained on 5000 samples, where a smooth dynamic was learned that closely resembled the true underlying process. In conclusion, MultiNODEs can already learn longitudinal dynamics based on only a few data points, however, they tend to underfit under these circumstances and benefit from larger sample sizes.

Adding an increasing noise level to the SIR training data revealed that MultiNODEs remain very robust **(Figure 6 C)**. Only when introducing 100% of the maximal encountered value as additional noise, a clear deviation from the underlying true model could be observed.

## Discussion

In this work, we presented MultiNODEs, a hybrid AI approach to generate synthetic patient-level datasets. MultiNODEs are specifically designed to consider the characteristics of clinical studies, as extends its predecessor, the Neural ODEs, and enables the application of the latent ODE system to multimodal datasets comprising both time-dependent and static variables with values missing not at random. MultiNODEs learn a latent, continuous time trajectory from observed data. This concept fits well to processes like disease progression, where relevant observations (e.g., biomarkers and disease symptoms) only indirectly mimic the true, underlying disease mechanism. Consequently, MultiNODEs are well suited for an application to heterogeneous datasets holding complex signals as encountered, for example, in biomedical research.

Our evaluations showed that MultiNODEs successfully generated complex, synthetic medical datasets that accurately reproduced the characteristics of their real-world counterparts. In a direct comparison MultiNODEs’ outperformed the state-of-the-art VAMBN approach, most notably with respect to the integrity of the correlation structure. This finding implies that the single data instances generated using MultiNODEs exhibit more realistic properties and that the real data characteristics are not only reproduced on population-level. Out of MultiNODEs two generative methods, the posterior sampling expectedly led to more realistic synthetic patients, however, generating from the prior distribution comes with the benefit that the model itself can be shared and used for data generation without needing any real datapoints in the process.

Machine learning classifiers that discriminated between real healthy controls and diseased subjects showed almost equal performance when trained on data from synthetic and real diseased subjects, respectively. Here, we only observed small deviations from the performance on real data for the NACC dataset, where classifiers trained and tested on synthetic patients and real healthy controls within a cross-validation setting showed a slightly increased performance to those trained on real data. Interestingly, at the same time we found a lower prediction performance compared to real data when we trained on synthetic subjects and evaluated on the real data. A possible explanation is that synthetic data can contain noise that is introduced during the generation of synthetic data points (for example through overestimated correlations between variables). Therefore, synthetically generated diseased patients are better discriminated against real healthy controls than real diseased patients. At the same time this situation leads to the fact that a classifier trained on synthetic data (synthetic patients as well as healthy controls) shows a slightly lower prediction performance on real data compared to a classifier trained entirely on real data. Altogether our results demonstrate that synthetically generated subjects share patterns of real patients, but they are not completely identical.

Besides the reproduction of marginal distributions and synthesis of realistic data instances, MultiNODEs most prominent strength lies in the generation of smooth longitudinal data. The latent ODE system allows MultiNODEs to learn dynamics which are continuous in time and cover the unobserved time intervals of real-world data. Here, both the prior and posterior sampling approach resulted in synthetic trajectories that obey real variables’ dynamics.

Furthermore, the time-continuous generative capabilities of MultiNODEs create opportunities to fill gaps in the real data through interpolation and go beyond the observation time by extrapolating the longitudinal dynamics. Hence, MultiNODEs could be used to support the design of longitudinal clinical studies, in which the maximum observation period as well as visit frequency are always crucial decisions to make. Here, the question of how patients might develop between two visits or after the last one determines the optimal follow-up time, to demonstrate, for example, the most significant treatment effect. Furthermore, synthetic disease trajectories generated based on data from one clinical study can be compared to those generated based on other studies, even if the visit intervals employed in the real studies were not identical.

Our benchmark experiments on the simulated SIR model data demonstrated that MultiNODEs are applicable under a variety of different data settings. While the general trends of a data underlying process could already be learned from a relatively limited dataset, similar to any machine learning task, the accuracy and trustworthiness of the model critically depends on the available data. Especially for complex, nonlinear processes, a sufficiently high observation frequency should be considered. Here, the position of the observation time-points relative to the true underlying process is crucial for MultiNODEs to accurately learn nonlinear dynamics. The sample size of the training data mainly impacts how well MultiNODEs fitted the data dynamics and we observed that lower sample sizes can lead to underfitting and rather rigid ODE systems. On the other hand, only severe noise levels led to a model deviation from the true data-underlying process and thus, with respect to noise, MultiNODEs proved to be highly robust. In conclusion, MultiNODEs’ requirements towards the training data ultimately depend on the complexity of the data underlying process, whereas the learning of more complex processes requires more frequent observations and larger sample size, while more linear systems can already be learned from rather limited datasets.

One limitation of MultiNODEs in their current form only allow static categorical variables. This is, because the variational encoder for longitudinal data maps trajectories to a latent Gaussian distribution. Sampling from this distribution (even, if conditioned on the distribution of the static data) and decoding will result into real valued features rather than categorical ones. In future work, we will thus explore whether a recurrent version of the HI-VAE encoder can be used instead of a recurrent variational LSTM encoder.

Additionally, MultiNODEs are sensitive to several hyperparameters that should be optimized for optimal performance. The training process and all relevant hyperparameters are explained in the Method section.

Synthetic data generated using models trained on sensitive personal information, can bear a risk of information disclosure (e.g. attribute disclosure or dataset membership disclosure), if an attacker has information about properties of real patients that are similar to a synthetic subject. Therefore, before synthetic data are distributed, it must be assured that the probability of private information disclosure remains within task-appropriate boundaries [23]. Disclosure risk often stands in a direct trade-off with data utility and a sensible compromise should be taken balancing the two according to the application in question. Several approaches are described in the literature that can reduce the risk of information disclosure [24], one of which is based on the concept of differential privacy [4]. MultiNODEs themselves provide a way to tune the deviation from the real data when sampling from the posterior distribution by changing the amount of noise injected in the latent space.

We would like to mention that a rigorous quantification of the re-identification risk is a non-trivial and challenging task for its own requiring several assumptions and is thus beyond the scope of this paper.

## Methods

### Application case datasets

Both datasets, namely PPMI and NACC, are well known staples in their respective fields and can be accessed after successful data access applications. For PPMI see https://www.ppmi-info.org/. For NACC we refer to https://naccdata.org/. More details on the investigated variables are presented in the Supplementary Material.

Both studies retrieved informed consent from their participants for data collection and sharing and followed the declaration of Helsinki to ensure ethical data collection. Both studies got ethical approval by their respective review boards. We followed their employed regulations and thus did not seek further ethical approval, as we did not work with human participants ourselves.

### Neural ODEs (NODEs)

NODEs are a hybrid of neural networks and ODEs [10]. They can be seen as an extension of a ResNet [16], which does not rely on a discrete sequence of hidden layers, but on a continuous hidden dynamical system defined by an ordinary differential equation.

For 0 <*t* < *M* and *z*_0_ ∈ *R*^*D*^ the dynamics of the hidden layer of a NODE are given as Equation (2).

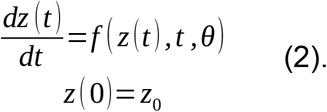

where *z* (0) may be interpreted as the first hidden layer and *z* (*T*) as the solution to the initial value problem at time point *T*. Importantly, *f* is a feed-forward neural network parameterized by *θ*.

### NODEs as generative latent time series models

As demonstrated by the authors in their publication, NODEs can be trained as a continuous time Variational Autoencoder. The basic idea is to learn the initial conditions *z*_0_ of the dynamical system in Equation (2) from observed time series data using a variational long-short term memory (LSTM) recurrent encoder [17]. Hence, Equation (2) now describes the dynamics of a latent system, resulting into a classical state-observation model. Accordingly, a feed-forward neural network decoder is required to project the solution of Equation (2) back to observed data at defined time points (**Supplementary Figure 10**).

Overall NODEs are trained at once by maximizing the Evidence Lower Variational Bound (ELBO): Let the training data be 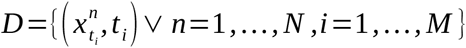, where *N* is the number of patients and 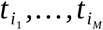 the observed time points / patient visits. That means 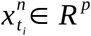 is the *p*-dimensional vector of measurements taken for the *n*-th patient at visit *t*_*i*_. The ELBO for NODEs is then given as Equation (3).

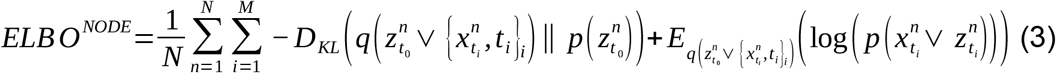

where 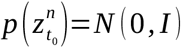, as usual. For details we refer to Chen *et al*. [10]

### Multi Modal Neural NODEs (MultiNODEs)

#### Handling missing values

To handle missing values (potentially not at random) in longitudinal clinical data we build on our previously published work, in which we introduced an imputation layer to implicitly estimate missing values during neural network training [14]: Let 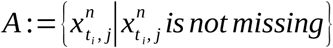, 1_*A*_ be the indicator function on set *A* with cardinality |*A*|. The imputation layer can be defined as a data transformation 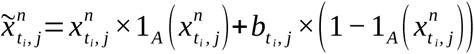, where parameters 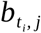 are trainable weights. That means missing values in a patient’s data vector 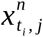 are replaced by 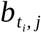. The accordingly completed data is subsequently mapped through a recurrent neural network encoder to a static, lower dimensional vector, which is interpreted as the initial condition of the latent ODE system (**Supplementary Figure 11**).

To learn parameters 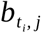 the NODEs’ loss function needs to be adapted. More specifically, we use the modified ELBO criterion presented in Equation (4).

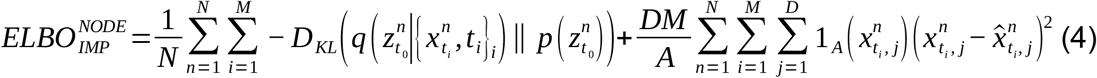

where 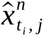 denotes the reconstructed data. Note that we only aim for reconstructing the observed data, but not the imputed one. Due to the layer-wise architecture of a neural network 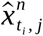 implicitly depends on 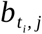.

In practice we initialize 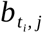 for neural network training as 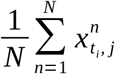

#### Dealing with multimodal data

In addition to implicit missing value imputation, the second main idea of MultiNODEs is to complement NODEs with a HI-VAE encoder [13] for static variables (**Supplementary Figure 11**). A HI-VAE is an extension of a Variational Autoencoder which can implicitly impute missing values via an input drop-out model and handle heterogeneous multimodal data, including categorical data and count data, via an accordingly factorized generative model. In addition, a HI-VAE uses a Gaussian Mixture Model (GMM) as a prior distribution rather than a single Gaussian. We refer to Nazabal *et al*. [13] for details.

The HI-VAE results in a lower dimensional latent representation *z*_*stat*_ of static variables, which can be used to augment the initial conditions 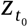 learned from time series data. Consequently, we arrive at the following formulation of the latent ODE system given in Equation (5).

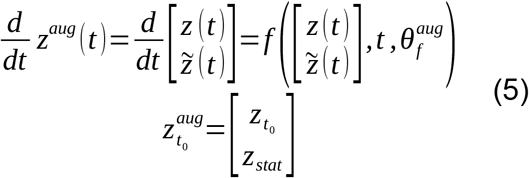

This approach resembles the Augmented Neural ODEs by Dupont *et al*. [18]. The main difference to our work is that in their work no additional features are added by the augmentation step, i.e. *z*_*stat*_ =0. According to Dupont et al. the purpose of Augmented Neural ODEs is to smoothen *f*, whereas we focus here on multimodal data integration.

For training MultiNODEs, we have to jointly consider 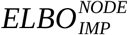 as well as *ELBO*^*HI -VAE*^. After bringing both quantities on a comparable numerical scale, we use a weighted sum as our final training objective (see Equation (6) and (7)):

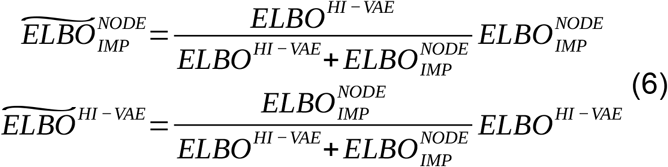

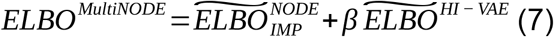

Where*β* is a tunable hyperparameter. Details about hyperparameter optimization are described in the Supplements.

#### Generating synthetic subjects

We tested two methods to generate synthetic subjects with MultiNODEs:

a. The first option is drawing a sample of latent static and longitudinal representations from the respective prior distributions 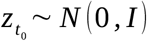 and *z*_*stat*_ ∼ *GMM* (*π*). To assure that interdependencies between static and longitudinal variables are conserved, we model their joint distribution *P* (*z*_*t* 0_, *z*_*stat*_) using a Bayesian network. This network contains three nodes (random variables) representing 1) the GMM mixture coefficients *π* for the static data used by the HI-VAE, 2) the latent static representations *Z*_*stat*_ =*GMM* (*π*), and 3) the latent longitudinal representations *Z*_*t* 0_=*N* (0, *I*), respectively. The network is constraint such that directed edges can only go from the *π* to *Z*_*stat*_ and from there to *Z*_*t* 0_. After randomly sampling a mixture component *s*_*i*_ from a multinomial distribution *multinom* (*π*), we can conditionally sample *z*_*stat*_ ∼ *Z* _*stat*_ ∨ *s*_*i*_ and finally *z*_*t* 0_ ∼ *Z*_*t* 0_ ∨ *Z*_*stat*_. Subsequently, we concatenate 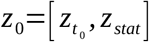 into a vector forming the initial conditions for the latent ODE system, solve the ODE system and decode the solution. We call this approach “prior sampling”.
b. A second option is to draw 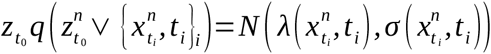 for the longitudinal data and 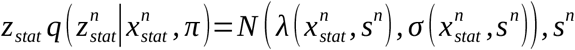 *Categorical* 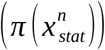 for the static data. That means we generate a blurred / noisy version of the original *n*-th patient. We call this approach “posterior sampling” and recommend this sampling procedure for data generation. In our experiments, we doubled the posterior variance during sampling because we found the synthetic data otherwise to lie too close to the real data. Tuning the added noise can provide one option to balance identification risk versus data utility.
c. Synthetic data can not only be generated for observed visits, but also for definable time points in between (interpolation) and after the end of study (extrapolation). This is possible because the latent ODE system is continuous in time.

#### Data preprocessing

Few steps are required to preprocess the clinical data before MultiNODEs can be applied. First, the data must be organized into a three-dimensional tensor of the shape samples × time points × variables for the longitudinal variables, and samples × variables for the static ones. Furthermore, the longitudinal variables are then transformed into a progression score by subtracting the baseline value and normalizing them by the standard deviation of this variable at baseline.

#### Calculating the relative error for correlation matrices

The relative error between correlation matrices is calculated as the norm of the matrix describing the difference between the real correlation matrix and synthetic data correlation matrix divided by the norm of the real correlation matrix.

## Data Availability

All data used in our manuscript are freely available for the purpose of (non-profit) research

https://www.ppmi-info.org/access-data-specimens/download-data/

https://naccdata.org/

## Code availability

The code for MultiNODEs is available at https://github.com/philippwendland/MultiNODEs.

## Data availability

The PPMI dataset is available under: https://www.ppmi-info.org/. The NACC data is available under: https://naccdata.org/. The data is shared by the data owners after successful application.

The data generated for this study can not be shared by the authors due to the signed data usage agreements with the data owners of the corresponding real data (ie PPMI and NACC).

## Acknowledgements

This project has received funding from the European Union’s Horizon 2020 research and innovation programme under grant agreement No. 826421, “TheVirtualBrain-Cloud” and from the Deutsche Forschungsgemeinschaft (DFG) funded project “NFDI4Health” (project number 442326535).

Data used in the preparation of this article were obtained from the Parkinson’s Progression Markers Initiative (PPMI) database (www.ppmi-info.org/data). For up-to-date information on the study, visit www.ppmi-info.org. PPMI – a public-private partnership – is funded by the Michael J. Fox Foundation for Parkinson’s Research and funding partners, including [list the full names of all of the PPMI funding partners found at www.ppmi-info.org/fundingpartners].

The NACC database is funded by NIA/NIH Grant U01 AG016976. NACC data are contributed by the NIA-funded ADCs: P30 AG019610 (PI Eric Reiman, MD), P30 AG013846 (PI Neil Kowall, MD), P30 AG062428-01 (PI James Leverenz, MD) P50 AG008702 (PI Scott Small, MD), P50 AG025688 (PI Allan Levey, MD, PhD), P50 AG047266 (PI Todd Golde, MD, PhD), P30 AG010133 (PI Andrew Saykin, PsyD), P50 AG005146 (PI Marilyn Albert, PhD), P30 AG062421-01 (PI Bradley Hyman, MD, PhD), P30 AG062422-01 (PI Ronald Petersen, MD, PhD), P50 AG005138 (PI Mary Sano, PhD), P30 AG008051 (PI Thomas Wisniewski, MD), P30 AG013854 (PI Robert Vassar, PhD), P30 AG008017 (PI Jeffrey Kaye, MD), P30 AG010161 (PI David Bennett, MD), P50 AG047366 (PI Victor Henderson, MD, MS), P30 AG010129 (PI Charles DeCarli, MD), P50 AG016573 (PI Frank LaFerla, PhD), P30 AG062429-01(PI James Brewer, MD, PhD), P50 AG023501 (PI Bruce Miller, MD), P30 AG035982 (PI Russell Swerdlow, MD), P30 AG028383 (PI Linda Van Eldik, PhD), P30 AG053760 (PI Henry Paulson, MD, PhD), P30 AG010124 (PI John Trojanowski, MD, PhD), P50 AG005133 (PI Oscar Lopez, MD), P50 AG005142 (PI Helena Chui, MD), P30 AG012300 (PI Roger Rosenberg, MD), P30 AG049638 (PI Suzanne Craft, PhD), P50 AG005136 (PI Thomas Grabowski, MD), P30 AG062715-01 (PI Sanjay Asthana, MD, FRCP), P50 AG005681 (PI John Morris, MD), P50 AG047270 (PI Stephen Strittmatter, MD, PhD).

## Author Contribution

CB and PW contributed equally to this work. HF, CB, PW, and MK conceived the project. PW implemented the method. PW, MGF, and MS performed the experiments. CB and HF wrote the manuscript. PW, MK, and MS revised the manuscript. CB and HF supervised the work.

