## Supplementary Material for "Generation of realistic synthetic data using multimodal neural ordinary differential equations"

### Supplementary Results

**Supplementary Table 1:** Summary statistic of marginal distributions shown in Figure 2 in the main text.

| | Mean (SD) | 25 % | Median | 75% | JS-Div. |
|---|---|---|---|---|---|
| **SCOPA month 12** | | | | | |
| Real | 0.28 (1.11) | -0.52 | 0.12 | 0.92 | - |
| Posterior | 0.27 (0.94) | -0.42 | 0.18 | 0.76 | 0.01 |
| VAMBN | 0.31 (1.08) | -0.42 | 0.31 | 0.99 | 0.02 |
| Prior | 0.41 (0.8) | -0.06 | 0.46 | 0.89 | 0.03 |
| **UPDRS2 month 24** | | | | | |
| Real | 0.73 (1.34) | -0.15 | 0.59 | 1.33 | - |
| Posterior | 0.66 (1.23) | -0.17 | 0.41 | 1.37 | 0.01 |
| VAMBN | 0.7 (1.17) | -0.13 | 0.66 | 1.52 | 0.02 |
| Prior | 0.87 (1.17) | 0.06 | 0.88 | 1.64 | 0.03 |
| **A-beta 42 static** | | | | | |
| Real | 0.0 (1.0) | -0.65 | -0.05 | 0.55 | - |

| | | | | | |
|---|---|---|---|---|---|
| Posterior | 0.01 (1.2) | -0.71 | -0.11 | 0.54 | 0.01 |
| VAMBN | 0.02 (0.93) | -0.65 | 0.03 | 0.6 | 0.01 |
| Prior | -069 (3.72) | -1.65 | -0.52 | 0.77 | 0.01 |

### Additional figures for PPMI experiments

To provide an overview about more variables, we present additional plots equivalent to those in the main text below. Plots for all variables, including relevant summary statistics like those presented in Supplementary Table 1, can be found under: https://github.com/philippwendland/MultiNODEs

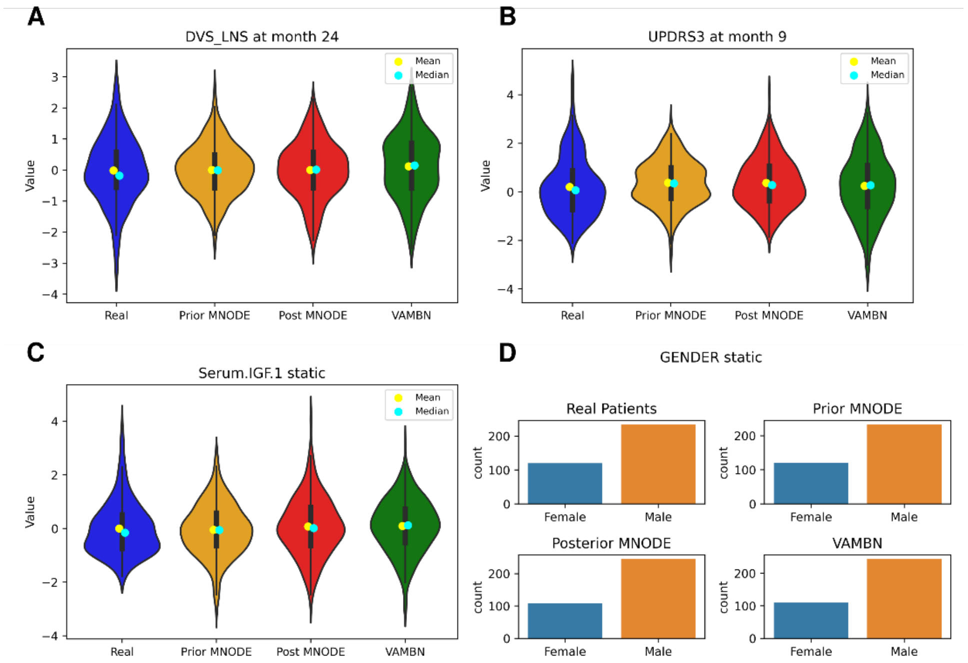


**Supplementary Figure 1:** Marginal distributions encountered in the generated synthetic data and real data of PPMI. Comprises additional examples corresponding to Figure 2 in the main text. Similar figures for all variables can be found at: https://github.com/philippwendland/MultiNODEs/tree/main/Plots/PPMI/Synthetic_Data_Generation/Static_violin

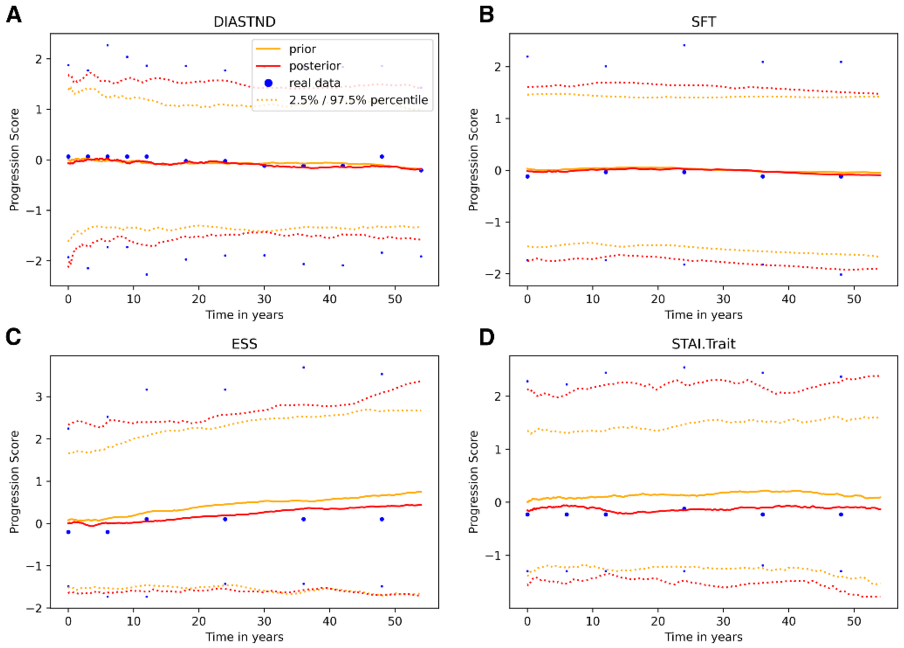


**Supplementary Figure 2:** Comparison of median trajectories including 2.5% / 97.5% quantiles of longitudinal variables from synthetic and real PPMI data. Comprises additional examples corresponding to Figure 3 in the main text. Similar figures for all variables can be found under: https://github.com/philippwendland/MultiNODEs/tree/main/Plots/PPMI/Synthetic_Data_Generation/Generated_Median.

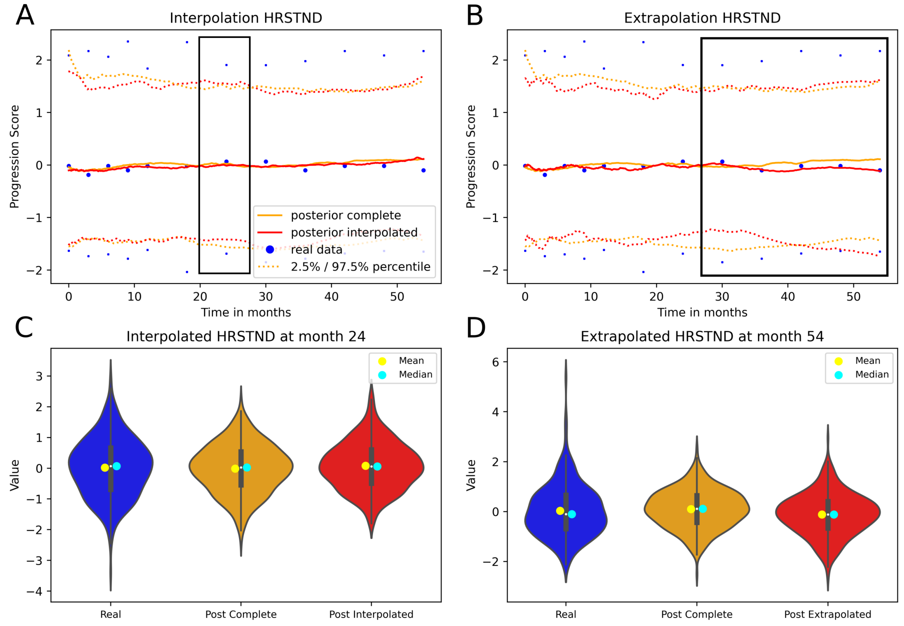


**Supplementary Figure 3:** Time-continuous interpolation and extrapolation of exemplary PPMI variables. The black box indicates the interpolated and extrapolated sections. Comprises additional examples corresponding to Figure 3 in the main text. Similar figures for all variables can be found under:

https://github.com/philippwendland/MultiNODEs/tree/main/Plots/PPMI. **A,** interpolation of the HRSTND variable at month 24. **B,** extrapolation of the last two assessments of the variable. **C,** marginal distribution of the interpolated values at visit 24. **D,** marginal distribution of the extrapolated values at month 48.

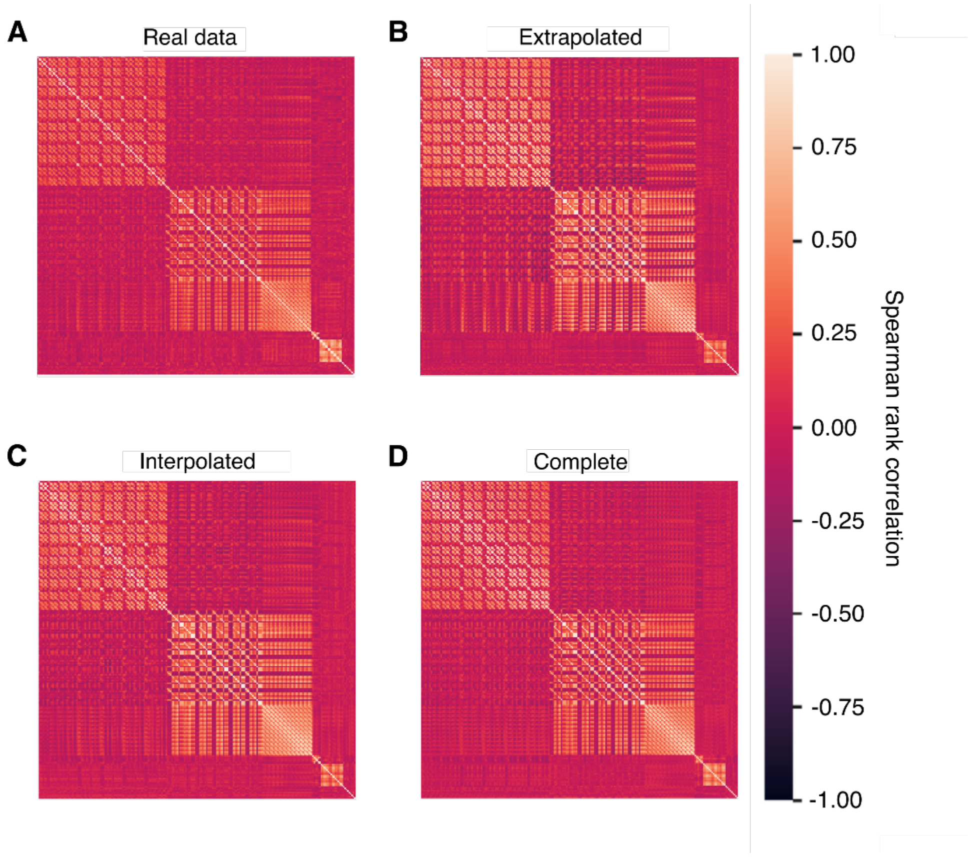


**Supplementary Figure 4:** Correlation structure of the interpolated and extrapolated PPMI data (i.e., generated using only parts of the real data) in comparison to the correlation structure of the real data and that of synthetic data generated based on the complete real data.

### NACC results

Below, we present the results based on the NACC dataset. All displayed results originate from experiments that were equivalent to those presented in the main text for the PPMI data. Figures for all experiments and all variables can be found under https://github.com/philippwendland/MultiNODEs/tree/main/Plots/NACC.

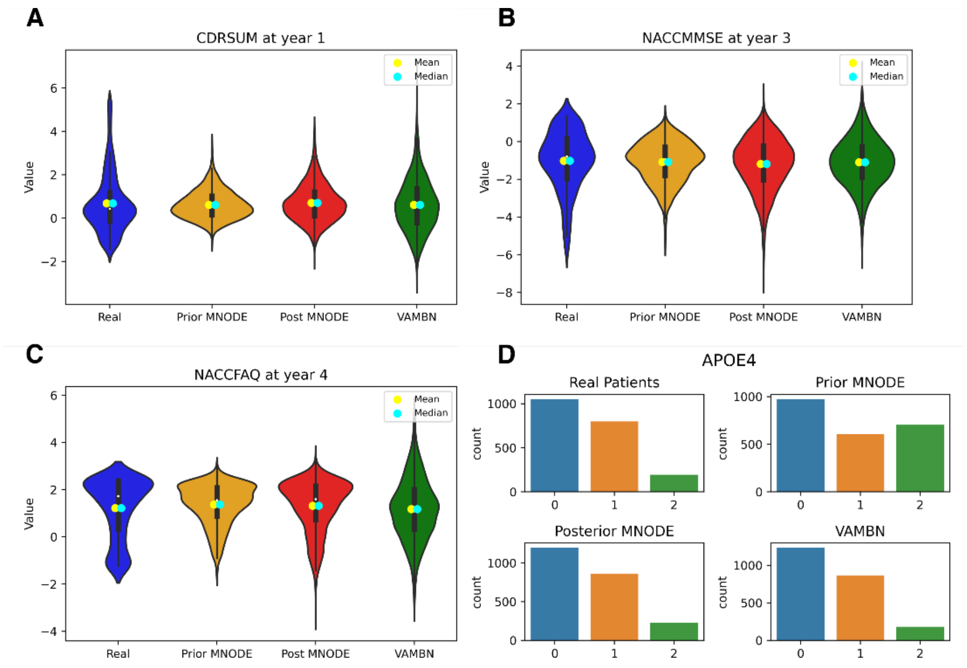


**Supplementary Figure 5:** Marginal distributions of real and synthesized data for multiple variables of the NACC data. Mean, standard deviation and KL-Divergence for the displayed variables can be found and similar figures for all NACC variables can be found under: https://github.com/philippwendland/Multimodal_Neural_ODEs/tree/main/Plots/NACC.

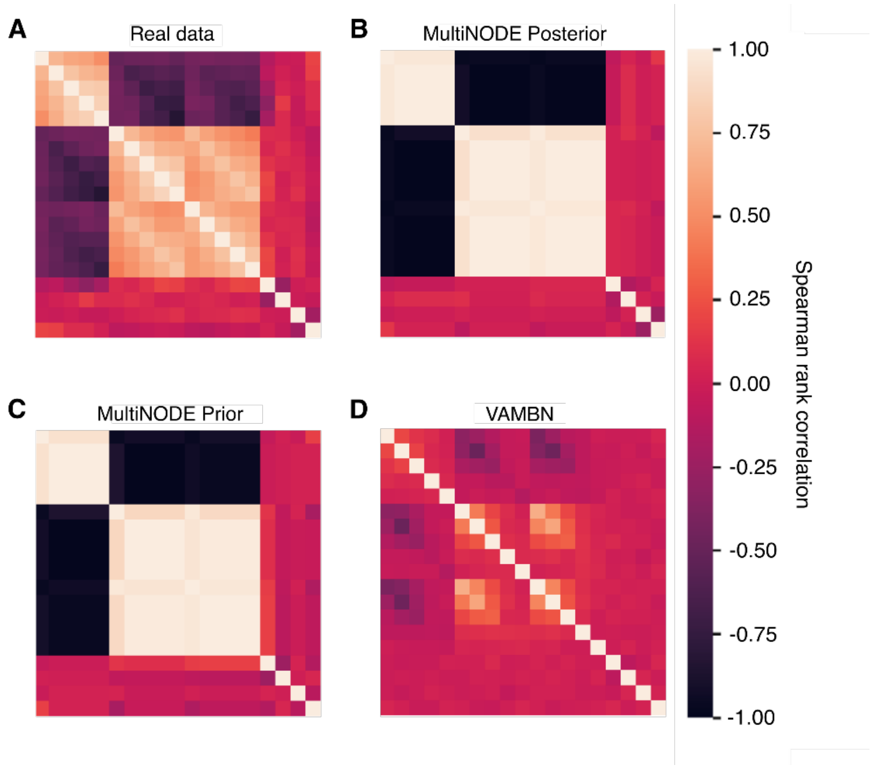

**Supplementary Figure 6:** Correlation structure of real and synthetic NACC data expressed as spearman rank correlation coefficients. **A,** real data. **B,** posterior sampling from MultiNODEs. **C,** prior sampling from MultiNODEs. **D,** VAMBN generated data.

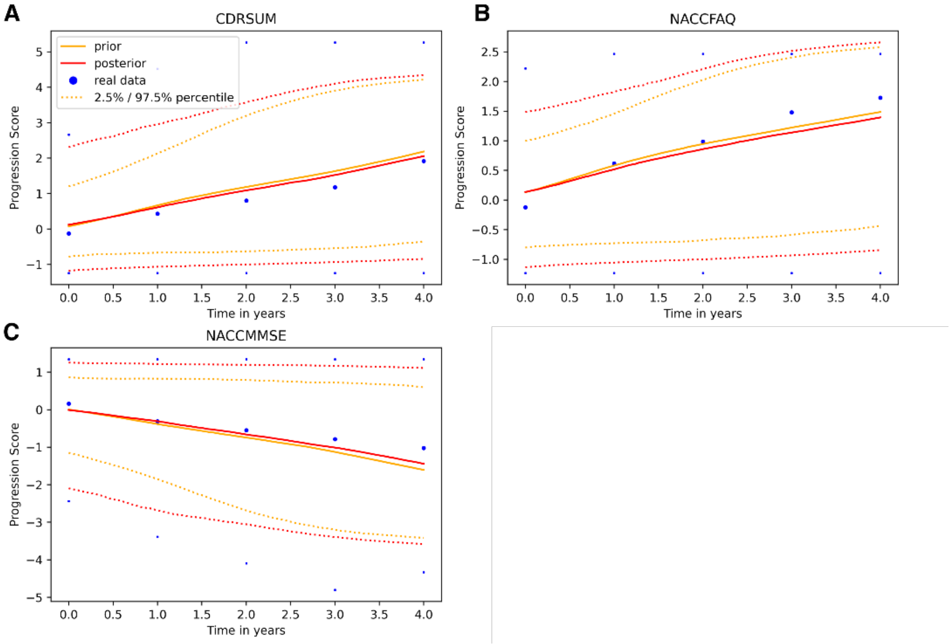


**Supplementary Figure 7:** Comparison of median trajectories including the 2.5% / 97.5% quantiles of longitudinal variables from synthetic and real NACC data. **A, B, C,** depict different longitudinal variables from the NACC dataset.

Interpolation of the NACC data was performed for year 2. For extrapolation, MultiNODEs were trained on data up to year 2 and the values for years 3 and 4 were extrapolated. Exemplary results are shown in **Supplementary Figure 8**. We observed that the mean JS-divergence calculated across all variables between the interpolated data and the real data was slightly higher (0.064 ± 0.054) than that of the real data and the synthetic data generated after training MultiNODEs on the complete trajectory (0.049 ± 0.026).

As in the interpolation setting, we again compared the average JS-divergence between the extrapolated data and the real data with that between the real data and synthetic data that were generated after training MultiNODEs on the complete trajectory. As expected, we could see a larger difference between the JS-divergences compared to the interpolation setting with 0.065 ± 0.024 for the extrapolated data and 0.022 ± 0.009 for the synthetic data based on the complete trajectory.

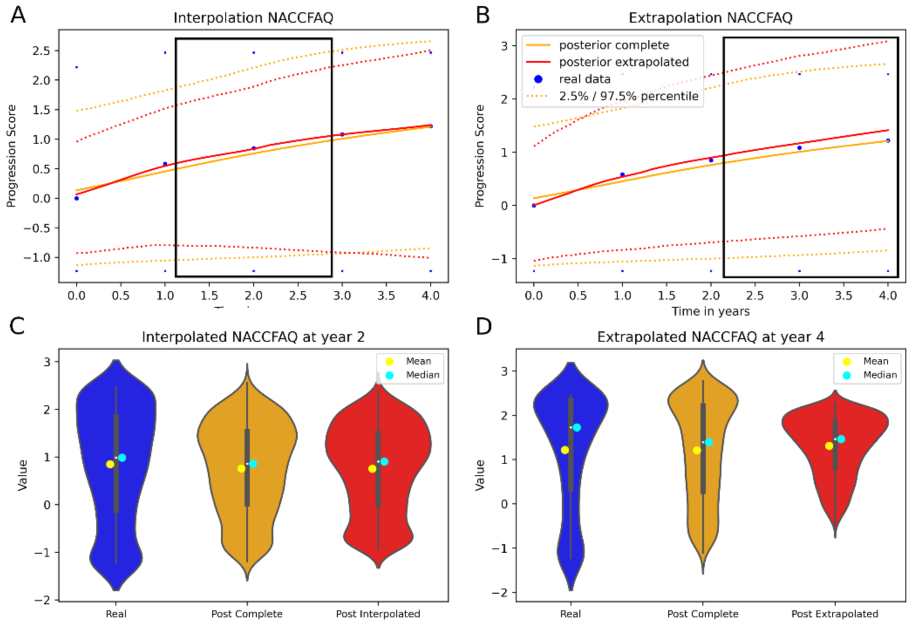


**Supplementary Figure 8:** Time-continuous interpolation and extrapolation of NACC variables. The black box indicates the interpolated and extrapolated sections. Similar figures for all variables can be found under: https://github.com/philippwendland/MultiNODEs/tree/main/Plots/NACC. **A,** interpolation of the FAQ variable at year 2. **B,** extrapolation of the last two assessments of the FAQ variable. **C,** distribution of the interpolated values for FAQ at year 2. **D,** distribution of the extrapolated values for FAQ at year 4.

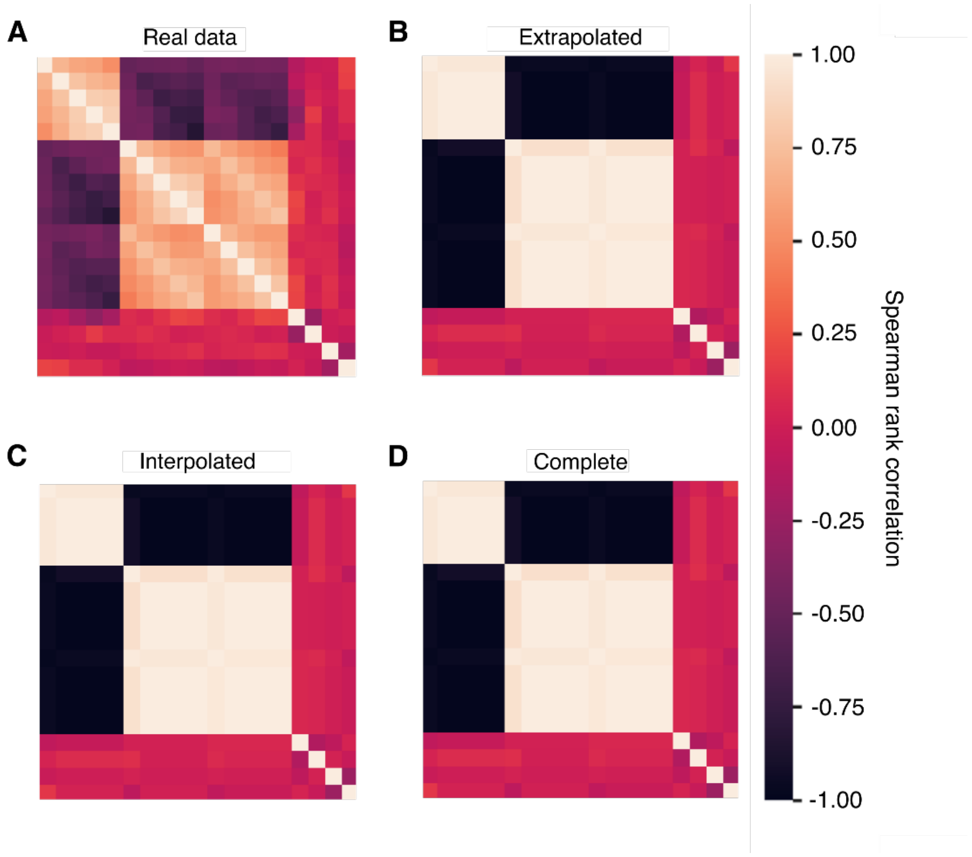


**Supplementary Figure 9:** Correlation structure of the interpolated and extrapolated NACC data (i.e., generated using only parts of the real data) in comparison to the correlation structure of the real data and that of synthetic data generated based on the complete real data.

### Supplementary Methods

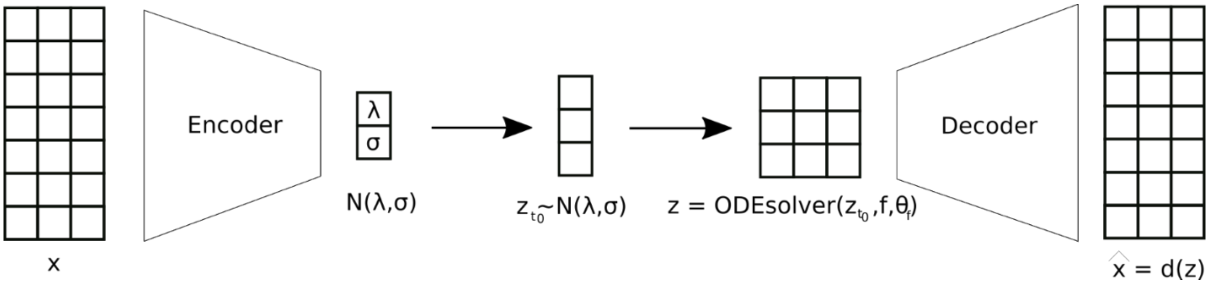


**Supplementary Figure 10:** Schema of a Neural ODE trained as a generative latent time series model

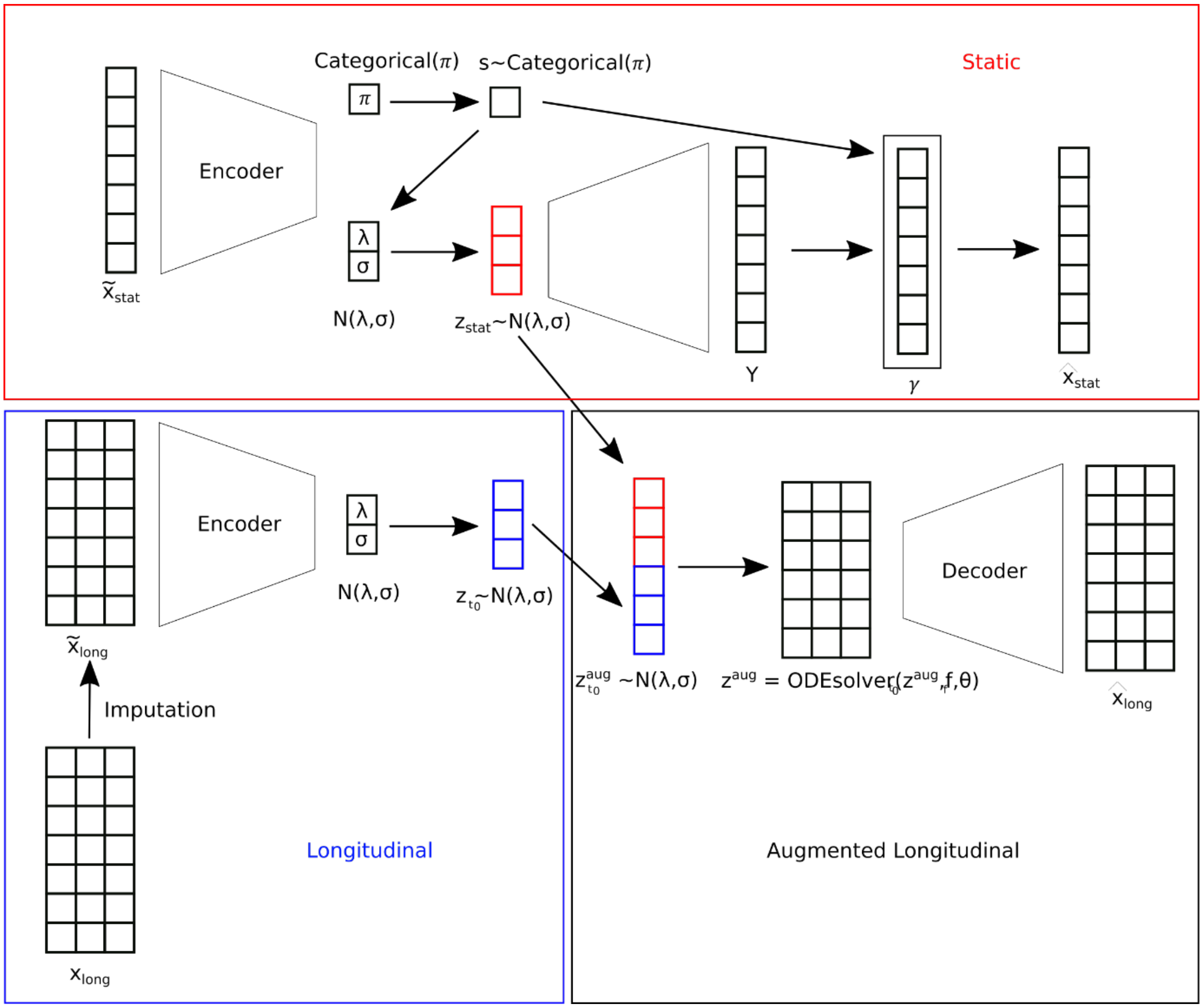


**Supplementary Figure 11:** Architecture of the MultiNODEs.

### Hyperparameter optimization

To apply MultiNODEs (and the VAMBN) to data, it is necessary to perform hyperparameter optimization. In this work, we used a Bayesian hyperparameter optimization [1] as implemented in the Optuna package [2]. We used the default settings employing a Tree of Parzen Estimator.

The target function to minimize was the weighted Mean Squared Error of the longitudinal data as reconstruction loss validated by a 5-fold-cross-validation. We used early stopping in every fold with a delta of 0 and a patience of 50 epochs. To determine the optimal number of epochs for the final model, we computed the average number of epochs across the folds. For the SIR model experiments, a simple train-test split was performed instead of a cross-validation due to the large sample sizes of the simulated data. The following hyperparameters were considered (the final hyperparameter configurations per model are presented at the end of this document):

| | |
|---|---|
| Learning rate | $[10^{-4}, 10^{-2}]$ |
| Number of epochs | {100, 200, …, 4000} |
| Batch size as fraction of the number of patients | [0, 1] |
| Encoder for longitudinal variables | Elman-network or LSTM |
| Number of hidden units of the encoder in percentage of the product of number of time points and number of longitudinal variables | [0, 1] |
| In the case of an Elman-network as an encoder: Activation function of the hidden layers | Tanh, ReLU or identity function |
| Dimension of initial condition (ignoring static features) of the latent ODE in percent of the number of longitudinal variables | [0, 4] |
| Number of hidden units of the feed-forward network representing the right-hand side of the latent ODE in percentage of the dimension of the initial conditions (ignoring static features) of the Neural latent ODE | [0, 10] |
| Steps to solve in the latent ODE | {1, 2, …, 8} |
| Activation function of the feed-forward network representing the right-hand side of the latent ODE | Tanh, ReLU, identity function |
| Number of hidden units in the decoder network, expressed as percentage of the product of number of timepoints and the number of longitudinal variables | [0, 1] |
| Activation function of the decoder | Tanh, ReLU, identity function |
| Fraction of input drop-out units in the decoder | [0, 1] |
| Number of mixture components in the Gaussian Mixture Model | {1, 2, …, 10} |
| Dimension of the latent representation of the static variables | {1,2, …,10} |
| Weighting parameter in MultiNODE ELBO | [0, 2] |

Supplementary Figure 12 shows a hyperparameter optimization run on the simulated SIR data.

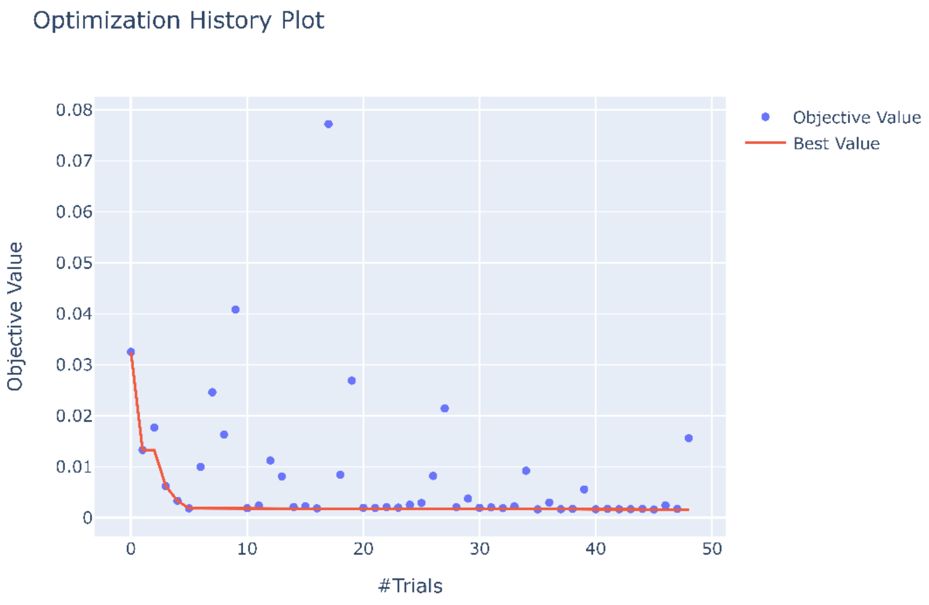


**Supplementary Figure 12:** Trials of a hyperparameter optimization run performed on the SIR data.

To train the VAMBN approach, it is again necessary to conduct a hyperparameter optimization. For every module of VAMBN, we performed a Bayesian hyperparameter optimization. We evaluated every configuration of hyperparameters with a 5-fold-cross-validation using the reconstruction loss (cross entropy) as a target function. The hyperparameters were the following:

| | |
|---|---|
| Learning rate | $[10^{-4}, 10^{-2}]$ |
| Mini-batch size | {16, 32} |
| Number of epochs | {200, …, 7000} |

### SIR Model

As parameters for the SIR model, we set T equal to 40, beta equal to 2, and gamma equal to 1. The population size was set to 1000.

**Classifier training**

We used the random forest implementation of sklearn with their default hyperparameters.  To impute missing values in the real data, we used the missForest iterative imputer of sklearn for a maximum of 100 iterations with 10 estimators.

**Supplementary Figure 13** shows the relative feature importance for the respective classifiers. Feature importance was calculated as decrease in classifier performance when the respective feature was excluded. For PPMI, the feature importance was accurately reflected in all synthetic classifiers. For NACC, we mainly saw lower importance of biological sex across all synthetic dataset-trained classifiers and an increase in importance for FAQ, a functional assessment of patients. Conclusively, the feature importance was quite stable between synthetic and real data-trained classifiers for both datasets.

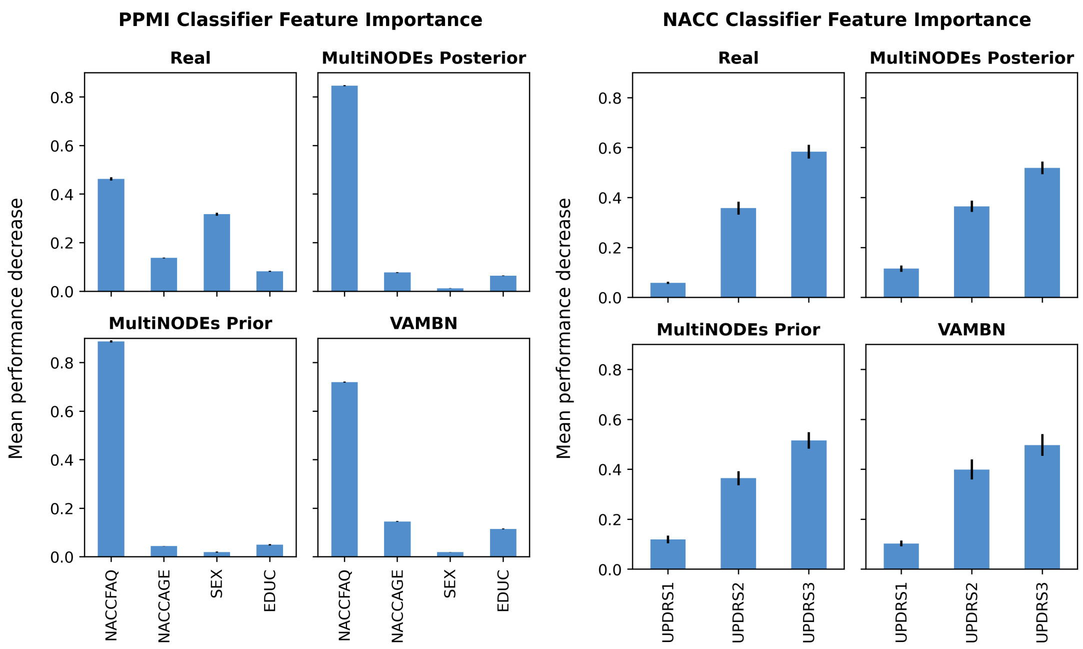


**Supplementary Figure 13:** Feature importance for predictors used to discriminate between real healthy control subjects and real and synthetic patients, respectively. Error bars depict the standard deviation calculated over 10 repeated runs of each classifier.

### Preprocessing of the PPMI data

To express the disease progression, we used z-scores normalized to a patient's baseline values. Let $x^n_{t_i, j_{long}}$ be the value of a longitudinal variable $j_{long}$ at a time point $t_i$ of patient $n$, $\mu_{t_{bl}, j_{long}}$ be the mean of a longitudinal variable at baseline and $\sigma_{t_{bl}, j_{long}}$ be the standard deviation of that variable at baseline. Then a z-score with respect to baseline can be defined as follows

$$\frac{x^n_{t_i, j_{long}} - \mu_{t_{bl}, j_{long}}}{\sigma_{t_{bl}, j_{long}}}$$

The static real valued variables were standardized, and the static categorical variables one-hot encoded.

To handle SNPs, we use CADD-filtered Impact Scores and Polygenetic Risk Scores. We receive odds ratios for the risk SNPs from the Phenome Wide Association Studies (PheWAS) catalog of genome-wide association studies (GWAS) [3]. To compute a polygenetic risk score for every patient, we sum up the odds ratio of the occurring risk SNPs per patient. The so-called Combined Annotation Dependent Depletion (CADD) values rate the maleficence of SNPs. To get CADD-filtered impact scores, we sum up the number of occurring risk SNPs over the recommended threshold of 15 per patient.

### List of Variables in PPMI

All longitudinal variables of the modules Medical History and UPDRS are measured at month 0, 3, 6, 9, 12, 18, 24, 30, 36, 42, 48, and 54. All longitudinal variables of the Non-Motor Module are measured at month 0, 12, 24, 36, and 48 (the following variables are measured at month 6th too: ESS, GDS, Quip, RBD, SCOPA, STAI.State, STAI.Trait, and STA). CSF-Alpha-Synuclein is measured at month 0, 6, 12, 24, 36.

| **Variable** | **Module** | **Name / Explanation** |
|---|---|---|
| WGTKG | Medical History | Weight in kilograms |
| HTCM | Medical History | Height in centimeters |
| TEMPC | Medical History | Temperature in Celsius |
| SYSSUP | Medical History | Systolic blood pressure (supine position) |
| DYSSUP | Medical History | Diastolic blood pressure (supine position) |
| HRSUP | Medical History | Heart frequency (supine position) |
| SYSTNP | Medical History | Systolic blood pressure (standing position) |
| DYSSTNP | Medical History | Diastolic blood pressure (standing position) |
| HRSTNP | Medical History | Heart frequency (standing position) |
| DVT-Total recall | Non-Motor | Subscore of Hopkins Verbal Learning Test |
| DVS-LNS | Non-Motor | Letter number sequencing |
| ESS | Non-Motor | Epworth‘s Sleepiness Scale |
| SCOPA | Non-Motor | Scales for outcomes in PD-autonomic |
| SFT | Non-Motor | Semantic fluency |
| STA | Non-Motor | State trait anxiety total score |
| STAI.State | Non-Motor | STAI – State subscore |
| STAI.Trait | Non-Motor | STAI – Trait subscore |

| | | |
|---|---|---|
| MDS-UPDRS 1 | UPDRS | Non-Motor values of daily living |
| MDS-UPDRS 2 | UPDRS | Motor values of daily living |
| MDS-UPDRS 3 | UPDRS | Motor examinations |
| CSF-Alpha-Synuclein | | Cerebrospinal fluid alpha-synuclein |

**Supplementary Table 2:** Longitudinal variables of the de-novo-patients (with modules of VAMBN)

| Variable | Module | Datatype | Name / explanation |
|---|---|---|---|
| A-beta42 | Biological | Numerical | Beta-amyloid 42 [39] |
| pTau | Biological | Numerical | Phospho-tau |
| Tau | Biological | Numerical | Total-tau |
| Tau/A-beta | Biological | Numerical | Total-tau/A42 |
| PTau/A-beta | Biological | Numerical | Phospho-tau/A-beta42 |
| pTau.Tau | Biological | Numerical | Phospho-tau/Total-tau |
| ALDH1A1..rep.1 | Biological | Numerical | Aldehyde dehydrogenase 1-family |
| ALDH1A1..rep.2 | Biological | Numerical | Aldehyde dehydrogenase 1-family |
| GAPDH..rep.1 | Biological | Numerical | Glyceraldehyde 3-phosphate dehydrogenase |
| GAPDH..rep.2 | Biological | Numerical | Glyceraldehyde 3-phosphate dehydrogenase |
| HSPA8..rep.1 | Biological | Numerical | Heat shock 70 kDa protein 8 |
| HSPA8..rep.2 | Biological | Numerical | Heat shock 70 kDa protein 8 |
| LAMB2..rep.1 | Biological | Numerical | Laminin subunit beta-2 |
| LAMB2..rep.2 | Biological | Numerical | Laminin subunit beta-2 |
| PGK1..rep.1 | Biological | Numerical | Phosphoglycerate kinase 1 |
| PGK1..rep.2 | Biological | Numerical | Phosphoglycerate kinase 1 |
| PSMC4..rep.1 | Biological | Numerical | 26S protease regulatory subunit 6B |
| PSMC4..rep.2 | Biological | Numerical | 26S protease regulatory subunit 6B |
| SKP1..rep.1 | Biological | Numerical | Guanine nucleotide exchange factor SPIKE 1 |
| SKP1..rep.2 | Biological | Numerical | Guanine nucleotide exchange factor SPIKE 1 |
| UBE2K..rep.1 | Biological | Numerical | Ubiquitin-conjugating enzyme E2 K |
| UBE2K..rep.2 | Biological | Numerical | Ubiquitin-conjugating enzyme E2 K |
| Serum.IGF.1 | Biological | Numerical | Insulin-like growth factor 1 |
| RAINDALS | Demographic | Categorical | Indians and indigenious americans |
| RAASIAN | Demographic | Categorical | Asians |
| RABLACK | Demographic | Categorical | Afroamericans |
| RAWHITE | Demographic | Categorical | White |
| RANOS | Demographic | Categorical | Not-specified ethnicity |
| EDUCYRS | Demographic | Numerical | Years of school education |
| HANDED | Demographic | Categorical | Handedness |
| Gender | Demographic | Categorical | Gender |
| BIOMOMPD | Family illness | Categorical | Biological mother has PD |
| BIODADPD | Family illness | Categorical | Biological father has PD |
| FULSIBPD | Family illness | Categorical | Biological siblings have PD |

| | | | |
|---|---|---|---|
| MAGPARPD | Family illness | Categorical | Maternal grandparents have PD |
| PAGPARPD | Family illness | Categorical | Paternal grandparents have PD |
| MATAUPD | Family illness | Categorical | Maternal aunts and uncles have PD |
| PATAUPD | Family illness | Categorical | Paternal aunts and uncles have PD |
| Imaging | | Categorical | DaTSCAN-scintigraphy |
| ENROLL AGE | | Numerical | Age at baseline |
| CADD filtered impact scores | | Numerical | Compare section… |
| Polygenetic risk scores | | Numerical | Compare section… |

**Supplementary Table 3:** Static variables of the PPMI de-novo PD patients.

#### NACC Variables

NACC's variables longitudinal variables were assessed annually over a period of up to 4 years.

| | | | |
|---|---|---|---|
| NACCMMSE | Longitudinal | Numerical | Mini-Mental State Examination |
| CDRSUM | Longitudinal | Numerical | Clinical Dementia Rating scale Sum of Boxes |
| NACCFAQ | Longitudinal | Numerical | Functional Activities Questionnaire |
| NACCNE4S | Static | Categorical | APOE E4 status |
| NACCAGE | Static | Numerical | Patient age at study baseline |
| SEX | Static | Categorical | Biological sex |
| EDUC | Static | Categorical | Years of education |

**Supplementary Table 4:** Variable description of the NACC study

#### Final model specifications

MultiNODEs implement several approaches to counteract overfitting to the training data. We implemented drop-out layers in the encoder, used cross-validation to tune hyperparameters, and the ELBO of the variational autoencoder framework represents another form of model regularization.

The models were optimized using the default implementation of pyTorch's Adam optimizer.

Below you find the chosen hyperparameters for all MultiNODEs used in the manuscript.

### PPMI Model Hyperparameters

| | |
|---|---|
| Learning rate | 0.0015680290621642827 |
| Number of epochs | 1900 |
| Batch size as fraction of the number of patients | 0.6400910825235765 |
| Encoder for longitudinal variables | Elman-network |
| Number of hidden units of the encoder in percentage of the product of number of time points and number of longitudinal variables | 0.49866725741363976 |
| In the case of an Elman-network as an encoder: Activation function of the hidden layers | Identity |
| Dimension of initial condition (ignoring static features) of the latent ODE in percent of the number of longitudinal variables | 1.8837462054054002 |
| Number of hidden units of the feed-forward network representing the right-hand side of the latent ODE in percent of the dimension of the initial conditions (ignoring static features) of the Neural latent ODE | 4.584466407303982 |
| Steps to solve in the latent ODE | 1 |
| Activation function of the feed-forward network representing the right-hand side of the latent ODE | Identity |
| Number of hidden units in the decoder network, expressed as percent of the product of number of timepoints and the number of longitudinal variables | 0.6974659448314735 |
| Activation function of the decoder | Identity |
| Fraction of input drop-out units in the decoder | 0.4951111384939868 |
| Number of mixture components in the Gaussian Mixture Model | 6 |
| Dimension of the latent representation of the static variables | 3 |
| Weighting parameter in MultiNODE ELBO | 0.9675595125571386 |
| Number of trainable parameters | 129373 |

### NACC Model Hyperparameters

| | |
|---|---|
| Learning rate | 0.0233168 |
| Number of epochs | 141 |
| Batch size as fraction of the number of patients | 0.656582 |
| Encoder for longitudinal variables | LSTM |
| Number of hidden units of the encoder in percentage of the product of number of time points and number of longitudinal variables | 3.03908 |
| In the case of an Elman-network as an encoder: Activation function of the hidden layers | |
| Dimension of initial condition (ignoring static features) of the latent ODE in percent of the number of longitudinal variables | 1.43289 |

| | |
|---|---|
| Number of hidden units of the feed-forward network representing the right-hand side of the latent ODE in percent of the dimension of the initial conditions (ignoring static features) of the Neural latent ODE | 4.00991 |
| Steps to solve in the latent ODE | 3 |
| Activation function of the feed-forward network representing the right-hand side of the latent ODE | Identity |
| Number of hidden units in the decoder network, expressed as percent of the product of number of timepoints and the number of longitudinal variables | 4.09659 |
| Activation function of the decoder | ReLU |
| Fraction of input drop-out units in the decoder | 0.0378523 |
| Number of mixture components in the Gaussian Mixture Model | 14 |
| Dimension of the latent representation of the static variables | 8 |
| Weighting parameter in MultiNODE ELBO | 1.60768 |
| Number of trainable parameters | 22241 |

### SIR standard

| | |
|---|---|
| Learning rate | 0.00678686 |
| Number of epochs | 676 |
| Batch size as fraction of the number of patients | 0.367031 |
| Encoder for longitudinal variables | Elman-network |
| Number of hidden units of the encoder in percentage of the product of number of time points and number of longitudinal variables | 0.436512 |
| In the case of an Elman-network as an encoder: Activation function of the hidden layers | TanH |
| Dimension of initial condition (ignoring static features) of the latent ODE in percent of the number of longitudinal variables | 1.04597 |
| Number of hidden units of the feed-forward network representing the right-hand side of the latent ODE in percent of the dimension of the initial conditions (ignoring static features) of the Neural latent ODE | 3.79528 |
| Steps to solve in the latent ODE | 5 |
| Activation function of the feed-forward network representing the right-hand side of the latent ODE | Identity |
| Number of hidden units in the decoder network, expressed as percent of the product of number of timepoints and the number of longitudinal variables | 0.910399 |
| Activation function of the decoder | TanH |
| Fraction of input drop-out units in the decoder | 0.00180114 |
| Number of mixture components in the Gaussian Mixture Model | 2 |

| | |
|---|---|
| Dimension of the latent representation of the static variables | 6 |
| Weighting parameter in MultiNODE ELBO | 1.3888 |
| Number of trainable parameters | 2035 |

#### SIR n=100

| | |
|---|---|
| Learning rate | 0.00601775920584171 |
| Number of epochs | 579 |
| Batch size as fraction of the number of patients | 0.3567612219833565 |
| Encoder for longitudinal variables | Elman-network |
| Number of hidden units of the encoder in percentage of the product of number of time points and number of longitudinal variables | 0.7504019762037379 |
| In the case of an Elman-network as an encoder: Activation function of the hidden layers | ReLU |
| Dimension of initial condition (ignoring static features) of the latent ODE in percent of the number of longitudinal variables | 1.010561217834188 |
| Number of hidden units of the feed-forward network representing the right-hand side of the latent ODE in percent of the dimension of the initial conditions (ignoring static features) of the Neural latent ODE | 3.6777847577609126 |
| Steps to solve in the latent ODE | 4 |
| Activation function of the feed-forward network representing the right-hand side of the latent ODE | Identity |
| Number of hidden units in the decoder network, expressed as percent of the product of number of timepoints and the number of longitudinal variables | 0.9328043779498616 |
| Activation function of the decoder | ReLU |
| Fraction of input drop-out units in the decoder | 0.048294395239418614 |
| Number of mixture components in the Gaussian Mixture Model | 1 |
| Dimension of the latent representation of the static variables | 6 |
| Weighting parameter in MultiNODE ELBO | 0.6323358754903038 |
| Number of trainable parameters | 2670 |

#### SIR N=5000

| | |
|---|---|
| Learning rate | 0.00998745 |
| Number of epochs | 1513 |
| Batch size as fraction of the number of patients | 0.17319 |
| Encoder for longitudinal variables | Elman-network |
| Number of hidden units of the encoder in | 0.226659 |

| | |
|---|---|
| percentage of the product of number of time points and number of longitudinal variables | |
| In the case of an Elman-network as an encoder: Activation function of the hidden layers | Identity |
| Dimension of initial condition (ignoring static features) of the latent ODE in percent of the number of longitudinal variables | 0.196854 |
| Number of hidden units of the feed-forward network representing the right-hand side of the latent ODE in percent of the dimension of the initial conditions (ignoring static features) of the Neural latent ODE | 1.59344 |
| Steps to solve in the latent ODE | 1 |
| Activation function of the feed-forward network representing the right-hand side of the latent ODE | TanH |
| Number of hidden units in the decoder network, expressed as percent of the product of number of timepoints and the number of longitudinal variables | 0.904631 |
| Activation function of the decoder | Identity |
| Fraction of input drop-out units in the decoder | 0.0515771 |
| Number of mixture components in the Gaussian Mixture Model | 6 |
| Dimension of the latent representation of the static variables | 2 |
| Weighting parameter in MultiNODE ELBO | 1.35137 |
| Number of trainable parameters | 424 |

### SIR t=5

| | |
|---|---|
| Learning rate | 0.00828768 |
| Number of epochs | 546 |
| Batch size as fraction of the number of patients | 0.683492 |
| Encoder for longitudinal variables | Elman-network |
| Number of hidden units of the encoder in percentage of the product of number of time points and number of longitudinal variables | 0.649597 |
| In the case of an Elman-network as an encoder: Activation function of the hidden layers | TanH |
| Dimension of initial condition (ignoring static features) of the latent ODE in percent of the number of longitudinal variables | 0.479475 |
| Number of hidden units of the feed-forward network representing the right-hand side of the latent ODE in percent of the dimension of the initial conditions (ignoring static features) of the Neural latent ODE | 0.812782 |
| Steps to solve in the latent ODE | 5 |
| Activation function of the feed-forward network representing the right-hand side of the latent ODE | Identity |
| Number of hidden units in the decoder network, | 0.857999 |

| | |
|---|---|
| expressed as percent of the product of number of timepoints and the number of longitudinal variables | |
| Activation function of the decoder | TanH |
| Fraction of input drop-out units in the decoder | 0.00545472 |
| Number of mixture components in the Gaussian Mixture Model | 2 |
| Dimension of the latent representation of the static variables | 4 |
| Weighting parameter in MultiNODE ELBO | 0.633321 |
| Number of trainable parameters | 507 |

### SIR t=100

| | |
|---|---|
| Learning rate | 0.00715729 |
| Number of epochs | 1480 |
| Batch size as fraction of the number of patients | 0.276469 |
| Encoder for longitudinal variables | Elman-network |
| Number of hidden units of the encoder in percentage of the product of number of time points and number of longitudinal variables | 0.00971514 |
| In the case of an Elman-network as an encoder: Activation function of the hidden layers | Identity |
| Dimension of initial condition (ignoring static features) of the latent ODE in percent of the number of longitudinal variables | 2.69398 |
| Number of hidden units of the feed-forward network representing the right-hand side of the latent ODE in percent of the dimension of the initial conditions (ignoring static features) of the Neural latent ODE | 0.515954 |
| Steps to solve in the latent ODE | 3 |
| Activation function of the feed-forward network representing the right-hand side of the latent ODE | TanH |
| Number of hidden units in the decoder network, expressed as percent of the product of number of timepoints and the number of longitudinal variables | 0.400573 |
| Activation function of the decoder | ReLU |
| Fraction of input drop-out units in the decoder | 0.173605 |
| Number of mixture components in the Gaussian Mixture Model | 6 |
| Dimension of the latent representation of the static variables | 2 |
| Weighting parameter in MultiNODE ELBO | 1.19349 |
| Number of trainable parameters | 2520 |

### SIR noise 50%

| | |
|---|---|
| Learning rate | 0.00523903 |
| Number of epochs | 220 |
| Batch size as fraction of the number of patients | 0.831626 |
| Encoder for longitudinal variables | LSTM |
| Number of hidden units of the encoder in percentage of the product of number of time points and number of longitudinal variables | 0.496277 |
| In the case of an Elman-network as an encoder: Activation function of the hidden layers | |
| Dimension of initial condition (ignoring static features) of the latent ODE in percent of the number of longitudinal variables | 1.20947 |
| Number of hidden units of the feed-forward network representing the right-hand side of the latent ODE in percent of the dimension of the initial conditions (ignoring static features) of the Neural latent ODE | 3.545 |
| Steps to solve in the latent ODE | 2 |
| Activation function of the feed-forward network representing the right-hand side of the latent ODE | Identity |
| Number of hidden units in the decoder network, expressed as percent of the product of number of timepoints and the number of longitudinal variables | 0.932988 |
| Activation function of the decoder | TanH |
| Fraction of input drop-out units in the decoder | 0.0961177 |
| Number of mixture components in the Gaussian Mixture Model | 6 |
| Dimension of the latent representation of the static variables | 3 |
| Weighting parameter in MultiNODE ELBO | 0.312058 |
| Number of trainable parameters | 2497 |

### SIR noise 75%

| | |
|---|---|
| Learning rate | 0.00417377 |
| Number of epochs | 208 |
| Batch size as fraction of the number of patients | 0.99479 |
| Encoder for longitudinal variables | Elman-network |
| Number of hidden units of the encoder in percentage of the product of number of time points and number of longitudinal variables | 0.502702 |
| In the case of an Elman-network as an encoder: Activation function of the hidden layers | TanH |
| Dimension of initial condition (ignoring static features) of the latent ODE in percent of the number of longitudinal variables | 1.21916 |
| Number of hidden units of the feed-forward network representing the right-hand side of the latent ODE in percent of the dimension of the initial conditions (ignoring static features) of the | 3.1724 |

| | |
|---|---|
| Neural latent ODE | |
| Steps to solve in the latent ODE | 4 |
| Activation function of the feed-forward network representing the right-hand side of the latent ODE | TanH |
| Number of hidden units in the decoder network, expressed as percent of the product of number of timepoints and the number of longitudinal variables | 0.277881 |
| Activation function of the decoder | Identity |
| Fraction of input drop-out units in the decoder | 0.0178918 |
| Number of mixture components in the Gaussian Mixture Model | 2 |
| Dimension of the latent representation of the static variables | 3 |
| Weighting parameter in MultiNODE ELBO | 0.933758 |
| Number of trainable parameters | 208 |

### SIR noise 100%

| | |
|---|---|
| Learning rate | 0.00123073 |
| Number of epochs | 508 |
| Batch size as fraction of the number of patients | 0.199226 |
| Encoder for longitudinal variables | Elman-network |
| Number of hidden units of the encoder in percentage of the product of number of time points and number of longitudinal variables | 0.79226 |
| In the case of an Elman-network as an encoder: Activation function of the hidden layers | ReLU |
| Dimension of initial condition (ignoring static features) of the latent ODE in percent of the number of longitudinal variables | 3.05187 |
| Number of hidden units of the feed-forward network representing the right-hand side of the latent ODE in percent of the dimension of the initial conditions (ignoring static features) of the Neural latent ODE | 2.7973 |
| Steps to solve in the latent ODE | 2 |
| Activation function of the feed-forward network representing the right-hand side of the latent ODE | ReLU |
| Number of hidden units in the decoder network, expressed as percent of the product of number of timepoints and the number of longitudinal variables | 0.277881 |
| Activation function of the decoder | Identity |
| Fraction of input drop-out units in the decoder | 0.95484 |
| Number of mixture components in the Gaussian Mixture Model | 6 |
| Dimension of the latent representation of the static variables | 5 |
| Weighting parameter in MultiNODE ELBO | 1.20909 |

| | |
|---|---|
| Number of trainable parameters | 4133 |
